# The time between vaccination and infection impacts immunity against SARS-CoV-2 variants

**DOI:** 10.1101/2023.01.02.23284120

**Authors:** Timothy A. Bates, Hans C. Leier, Savannah K. McBride, Devin Schoen, Zoe L. Lyski, David X. Lee, William B. Messer, Marcel E. Curlin, Fikadu G. Tafesse

**Author notes:** Corresponding authors Fikadu G. Tafesse –, Marcel E. Curlin –, William B. Messer –.

## Abstract

As the COVID-19 pandemic continues, long-term immunity against SARS-CoV-2 will be globally important. Official weekly cases have not dropped below 2 million since September of 2020, and continued emergence of novel variants have created a moving target for our immune systems and public health alike. The temporal aspects of COVID-19 immunity, particularly from repeated vaccination and infection, are less well understood than short-term vaccine efficacy. In this study, we explore the impact of combined vaccination and infection, also known as hybrid immunity, and the timing thereof on the quality and quantity of antibodies produced by a cohort of 96 health care workers. We find robust neutralizing antibody responses among those with hybrid immunity against all variants, including Omicron BA.2, and we further found significantly improved neutralizing titers with longer vaccine-infection intervals up to 400 days. These results indicate that anti-SARS-CoV-2 antibody responses undergo continual maturation following primary exposure by either vaccination or infection for at least 400 days after last antigen exposure. We show that neutralizing antibody responses improved upon secondary boosting with greater impact seen after extended intervals. Our findings may also extend to booster vaccine doses, a critical consideration in future vaccine campaign strategies.

## Introduction

Since the emergence of severe acute respiratory syndrome coronavirus 2 (SARS-CoV-2) in late 2019, the coronavirus disease 2019 (COVID-19) pandemic has continued to expand and contract at regular intervals, and it remains an ongoing threat to global public health. As of August 2022, the number of officially recognized cases is approaching 600 million,^1^ and the true number of people with at least one previous infection is likely much higher with estimates upwards of 3.4 billion, 44% of the global population, even before the emergence of the Omicron variant.^2^ Due to ongoing transmission and the continued emergence of novel SARS-CoV-2 variants, it is likely that this number will continue to rise despite large-scale public health control efforts. Nevertheless, current vaccines have proven to be invaluable tools for protecting public health and have saved countless lives.

First generation lipid nanoparticle mRNA vaccines including Comirnaty (Pfizer-BioNTech, previously BNT162b2) and Spikevax (Moderna, previously mRNA-1273) became available in the United States in December, 2020, and to this day remain the most utilized vaccines in many parts of the world.^3^ These vaccines are both well established as providing temporary prevention of SARS-CoV-2 infection as well as longer-term protection from severe COVID-19 and death.^4,5^ The primary challenges faced by vaccination-based protection at this stage in the pandemic are antibody waning and the emergence of concern (VOCs).^6,7^ Additional vaccine boosters given months after initial vaccination have been shown to provide partial protection against novel variants including Omicron.^8,9^ However, the most protective immune responses are seen after a combination of vaccination and natural infection, also known as hybrid immunity.^10–13^

Several key variables influence the protective efficacy of SARS-CoV-2 immunity. The first is the mechanisms by which immunity is elicited, which may include natural infection or vaccination with any of the different vaccine types.^13,14^ The second is viral antigenic variation, which encompasses differences in the amino acid sequence and post-translational modification of viral antigens depending on which variant of SARS-CoV-2 the antigens were derived from.^15,16^ The third is timing between repeat exposures, including the interval between vaccine doses and the much less studied interval between vaccination and natural infection.^17–20^ Additionally, the length of time since last exposure can lead to waning immunity and decreased protection. However, the durability of responses from different exposure modes can vary greatly.^13,21,22^ Finally, other variables exist which have important implications for immunity including a person’s age, sex, and comorbidities, and certain therapeutic agents. Understanding the impact of these variables is key for risk-stratifying populations and guiding general vaccination strategies.

As the pandemic continues, separating these variables’ individual contributions to immunity becomes increasingly complex, particularly as global efforts to track infections lose momentum. Further, as SARS-CoV-2 transitions to a globally endemic virus, hybrid immunity from combined vaccination and natural infection will be the dominant form of immunity, and while hybrid immunity is currently the subject of intense focus, very little work has been done thus far to determine the impact of exposure timing on its development.

Here, we report results of studies of 2 cohorts: the first is comprised of individuals recovered from COVID-19 and paired infection naïve, vaccinated controls from whom serum samples were collected both before and after vaccination; the second cohort builds on our experience from the first cohort and includes vaccinated individuals with prior COVID-19, vaccinated individuals that then experienced breakthrough infection, and infection naïve vaccinated controls. The second cohort includes individuals with a wide range of intervals (35-404 days) between PCR-confirmed COVID-19 and vaccination. We utilized enzyme-linked immunosorbent assays (ELISA) and live-virus neutralization assays with the original SARS-CoV-2 (WA1) and the variants of concern (Alpha, Beta, Gamma, Delta, Omicron BA.1, and Omicron BA.2) to discern how the interval between vaccination and infection affects the resulting level of humoral immunity. We find that the magnitude, potency, and breadth of the hybrid immune response against variants continue to improve for at least 400 days. These results suggest that the primary immune response to either vaccination or natural infection continues developing for over a year after first exposure, in the absence of additional exposures and that boosting with the vaccine or infection leads to a hybrid immunity with dramatically improved antibody quantity and quality as measured by their capacity to recognize and neutralize emergent SARS-CoV-2 variants.

## Results

### A longitudinal cohort of vaccinees with previous COVID-19 displayed improved SARS-CoV-2 neutralization compared to vaccination alone

Between December 2020 and March 2021, we recruited 10 individuals that experienced PCR-confirmed COVID-19 prior to vaccination and collected blood samples before and after a standard two-dose BNT162b2 vaccine regimen (Table 1) and 20 age and sex matched with no self-reported history of prior COVID-19 infection, verified by negative nucleocapsid ELISA, and collected blood samples before and after vaccination. We then measured and compared serum neutralizing titers in for these two groups using a live virus focus reduction neutralization test (FRNT) (Figure 1A-B). Serum neutralizing titers increased for both groups pre- and post-vaccination and were significantly higher among those with prior infection compared with vaccination for all strains tested, including ancestral strain of SARS-CoV-2 (WA1) as well as the early VOCs Alpha, Beta, and Gamma (Figure 1C). These results suggested that hybrid immunity from the combination of vaccination and natural infection may result in meaningfully improved neutralizing serum antibody titers.

**Table 1:** Demographics.

|  |  | Pre/post vaccination<br>(longitudinal) |  |  | Post vaccine (cross-sectional) |  |  |  |  |
| --- | --- | --- | --- | --- | --- | --- | --- | --- | --- |
|  |  | All | Vaccine<br>only | Prior<br>infection | All | Vaccine only | Hybrid immunity |  |  |
|  |  |  |  |  |  |  | All | Prior<br>infection | Breakthro<br>ugh<br>infection |
| Cohort size | n | 30 | 20 | 10 | 66 | 20 | 46 | 23 | 23 |
| Age | Years - median<br>[range] | 39.5<br>[23-63] | 41.5<br>[25-63] | 36.5 [23-61] | 39.5<br>[23-73] | 39.5 [23-63] | 40 [23-73] | 47 [23-73] | 38 [24-63] |
| Sex | Male - n (%) | 10 (33) | 6 (30) | 4 (40) | 18 (27) | 3 (15) | 15 (33) | 10 (43) | 5 (22) |
|  | Female - n (%) | 20 (67) | 14 (70) | 6 (60) | 48 (73) | 17 (85) | 31 (67) | 13 (57) | 18 (78) |
| disease<br>severity | Asymptomatic -<br>n (%) | - | - | 2 (20) | - | - | 3 (7) | 3 (13) | 0 (0) |
|  | Mild - n (%) | - | - | 7 (70) | - | - | 39 (85) | 19 (83) | 20 (87) |
|  | Moderate - n<br>(%) | - | - | 1 (10) | - | - | 3 (7) | 1 (4) | 2 (9) |
| between<br>vaccine<br>doses | Days - median<br>[range] | 22 [20-32] | 21 [21-32] | 22 [20-25] | 21 [17-45] | 21 [21-25] | 21 [17-45] | 22 [18-45] | 21 [17-32] |
| exposure<br>interval* | Days - median<br>[range] | - | - | 98 [40-303] | - | - | 221 [35-404] | 299 [40-404] | 215 [35-238] |
| collection<br>interval** | Days - median<br>[range] | 17 [10-28] | 16 [10-25] | 18 [14-28] | 23 [10-53] | 19.5 [10-28] | 25.5 [10-53] | 25 [11-53] | 27 [10-49] |

**Figure 1:**
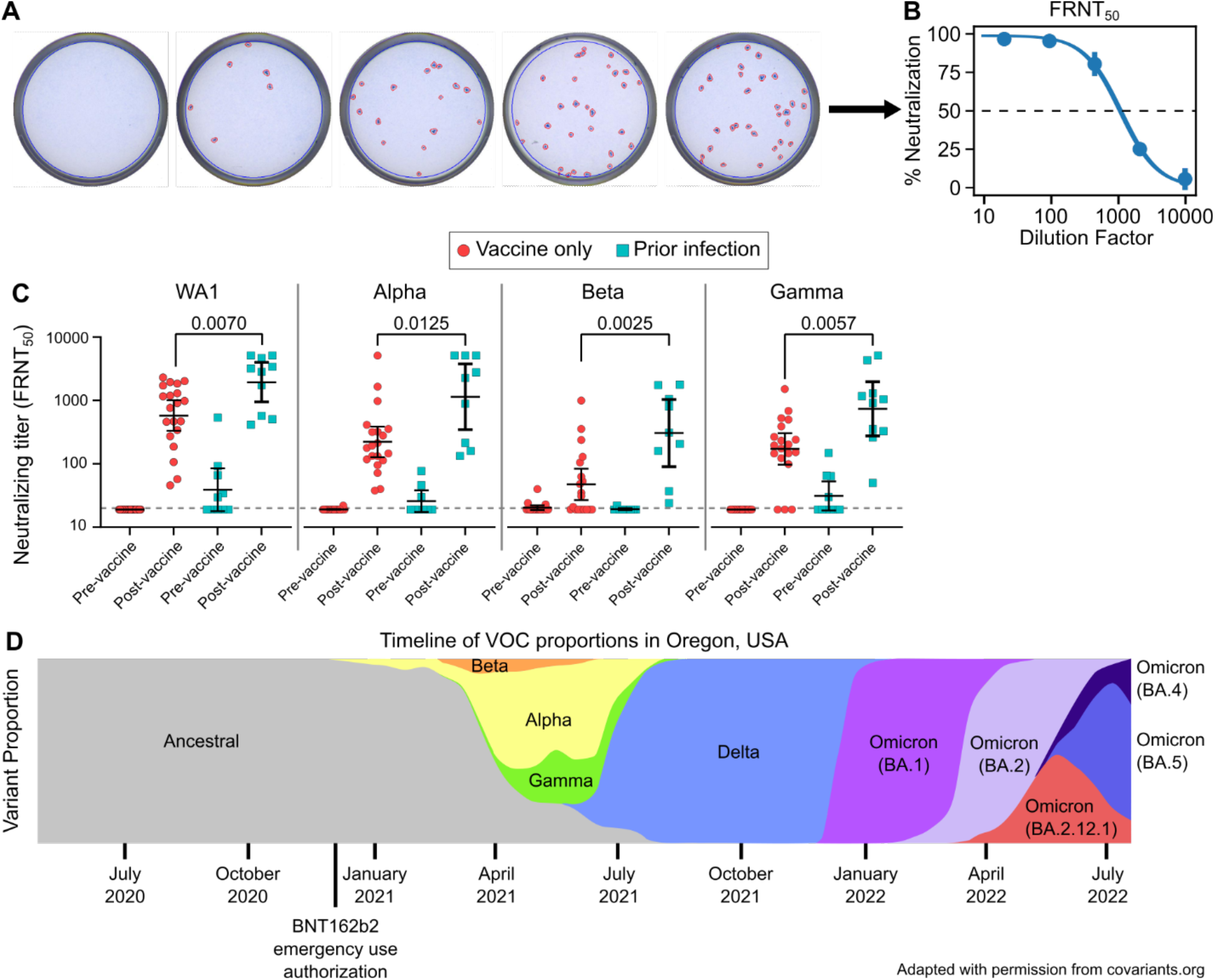
Longitudinal cohort of previously infected vaccinees shows improved variant neutralization compared to vaccination alone. Representative focus reduction neutralization test (FRNT) results showing wells infected with live SARS-CoV-2 with the addition of serially diluted serum which were stained and counted (**A**). Representative focus reduction neutralization curve showing the average neutralization of duplicates as a percent of no serum controls and fit to a dose-response curve to find the 50% neutralizing titer (FRNT_50_) (**B**). Live virus FRNT_50_ measurements against original SARS-CoV-2 (WA1) and the Alpha, Beta, and Gamma variants before and after vaccination (**C**). Timeline depicting the prevalence of impactful variants in the study location, Oregon, USA (**D**).^41^ Vaccine-only participants are represented by red circles and hybrid immune participants by cyan squares. Error bars represent the geometric mean with 95% confidence intervals. P values in C show the result of Mann-Whitney U tests. All P values are two-tailed and 0.05 was considered significant. For panel C, n=20 for the vaccine only group and n=10 for the prior infection group.

### A cross-sectional cohort of hybrid immune individuals including both prior infection and vaccine breakthrough

To more comprehensively study our initial results suggesting infection followed by vaccination elicited higher levels of SARS-CoV-2 specific antibodies compared to vaccination alone, we next expanded on our cohort by recruiting additional vaccinated persons with or without hybrid immunity due to previous COVID-19 (Table 1). This larger hybrid immune group included 23 individuals with PCR-confirmed infections prior to vaccination and 23 with vaccine breakthrough infections, as both vaccination/infection histories have been shown to provide similar levels of serological immunity.^11^ To assure a more uniform comparison, sera were collected less than 60 days following vaccination or PCR-confirmed breakthrough infection. The participants with infection prior to vaccination had all contracted COVID-19 during the pre-VOC era and are thus believed to have been infected with ancestral SARS-CoV-2 variants, while breakthrough cohort participants were recruited after the emergence of the VOCs, but prior to the Omicron era (Figure 1D). Using a subset of subjects for whom appropriate samples were available, viral sequences were obtained from 17 of 23 breakthrough participants with showing that the majority of infections were caused by the Alpha and Delta VOCs (Table 2).

**Table 2:** variants of infections.

|  | N | % |
| --- | --- | --- |
| **Prior infection** | 23 | 100 |
| Not sequenced* | 23 | 100 |
| **Breakthrough infection** | 23 | 100 |
| Alpha | 4 | 17 |
| Beta | 1 | 4 |
| Gamma | 2 | 9 |
| Delta | 10 | 43 |
| Not sequenced** | 6 | 26 |

### Elevated antibody levels and neutralizing titers with hybrid immunity

We next measured spike-specific antibody levels for our larger cohort with a series of ELISA experiments. Against purified RBD protein, total antigen-specific antibody levels increased 3.6-fold with hybrid immunity compared to vaccine only (Figure 2A). Class-specific ELISAs showed that this was primarily driven by improvements in IgG levels, which increased 3.7-fold (Figure 2B), while the less abundant IgA improved by 3.2-fold (Figure 2C), and IgM levels showed no significant difference between groups (Figure 2D). Total antibody levels against the full-length spike protein, which includes the entire S1 and S2 domains were also improved with hybrid immunity by 3.1-fold (Figure 2E).

**Figure 2:**
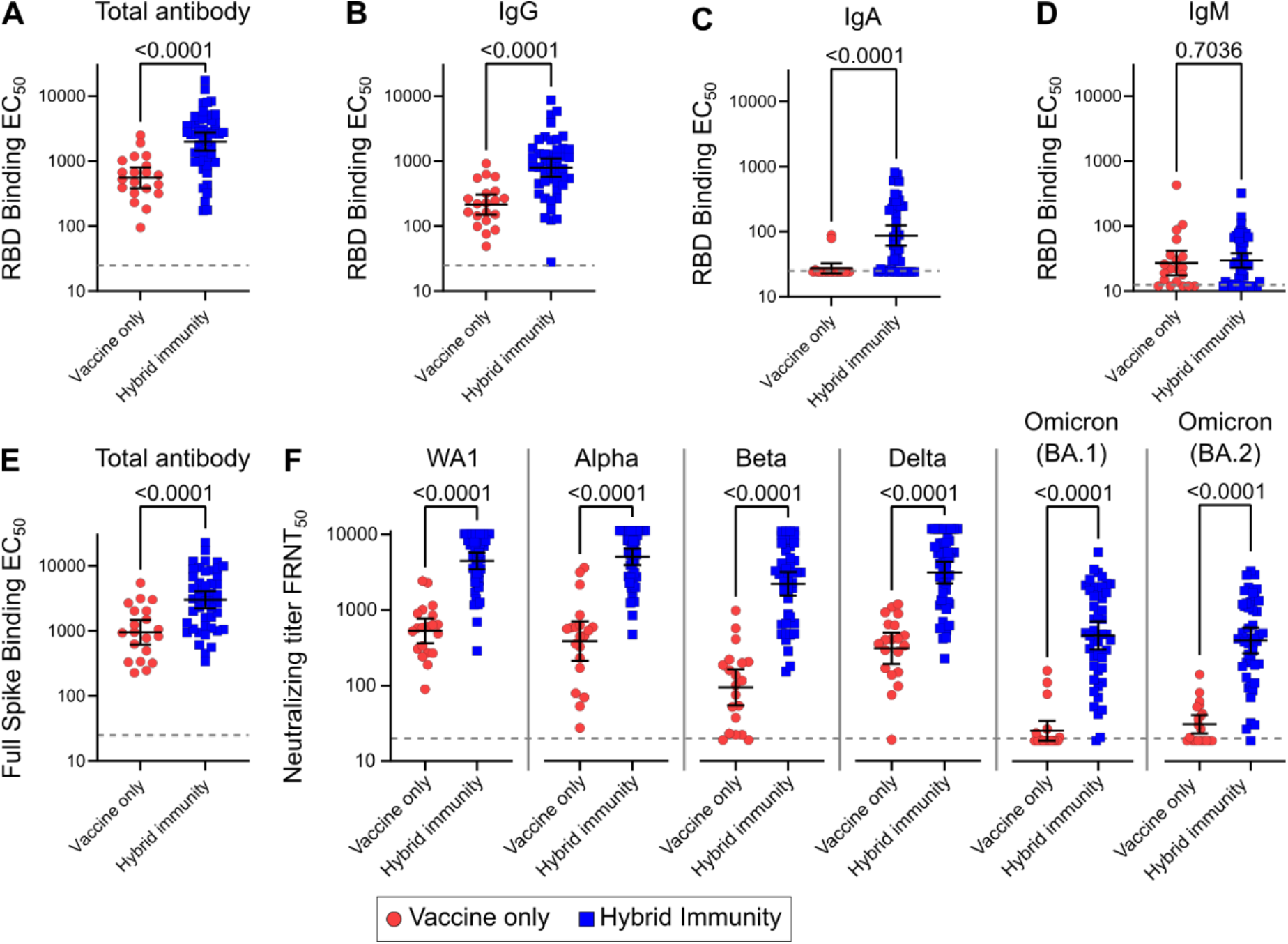
Cross-sectional cohort of individuals with hybrid immunity show improved antibody levels and variant neutralization. Levels of SARS-CoV-2 spike receptor binding domain (RBD)-specific total (IgG/A/M) antibody (**A**), IgG (**B**), IgA (**C**), and IgM (**D**). Levels of full-length spike-specific total antibody (**E**). Live virus FRNT_50_ measurements against original SARS-CoV-2 (WA1) and the Alpha, Beta, Delta, Omicron (BA.1), and Omicron (BA.2) variants (**F**). Vaccine only participants are represented by red circles and hybrid immune participants by blue squares. Error bars represent the geometric mean with 95% confidence intervals. P values in A-F show the result of Mann-Whitney U tests. All P values are two-tailed and 0.05 was considered significant. For panels A-F, n=20 for the vaccine only group and n=46 for the hybrid immunity group.

Similarly, neutralizing antibody titers against SARS-CoV-2 and every SARS-CoV-2 variant tested rose significantly in the hybrid immune group compared to vaccination alone (Figure 2F). Neutralizing titers improved by 8.4-fold against WA1, 12.5-fold against Alpha, 22.7-fold against Beta, 9.6-fold against Delta, 19.0-fold against Omicron BA.1, and 13.3-fold against Omicron BA.2. The largest fold-increases were seen against the most vaccine resistant variants, Beta and Omicron (BA.1 and BA.2). Further, it appears that these increases were not restricted to variants with which the cohort was experienced, as all samples were collected prior to the emergence of Omicron.

### Improved antibody quality among hybrid immune individuals

To assess the breadth of the neutralizing antibody response, we then looked at the relative ability to neutralize variants. This was measured by dividing the neutralizing titer for each variant by the neutralizing titer for WA1. For the Alpha and Beta variants, the hybrid immunity cohort showed greater cross-reactivity compared to vaccine only cohort and moves closer to equal neutralization compared to WA1 (Figure 3A and 3B). The difference does not appear to be as large for Delta (Figure 3C), while cross-neutralization against Omicron BA.1 and BA.2 did appear greater, but with cross-reactivity most marked among those with higher titers (Figure 3D and 3E). This is quantified by the cohort geometric mean variant cross-neutralization scores, which showed significantly greater cross-reactivity for all variants except for Delta (Figure 3F and supplemental figure 1).

**Figure 3:**
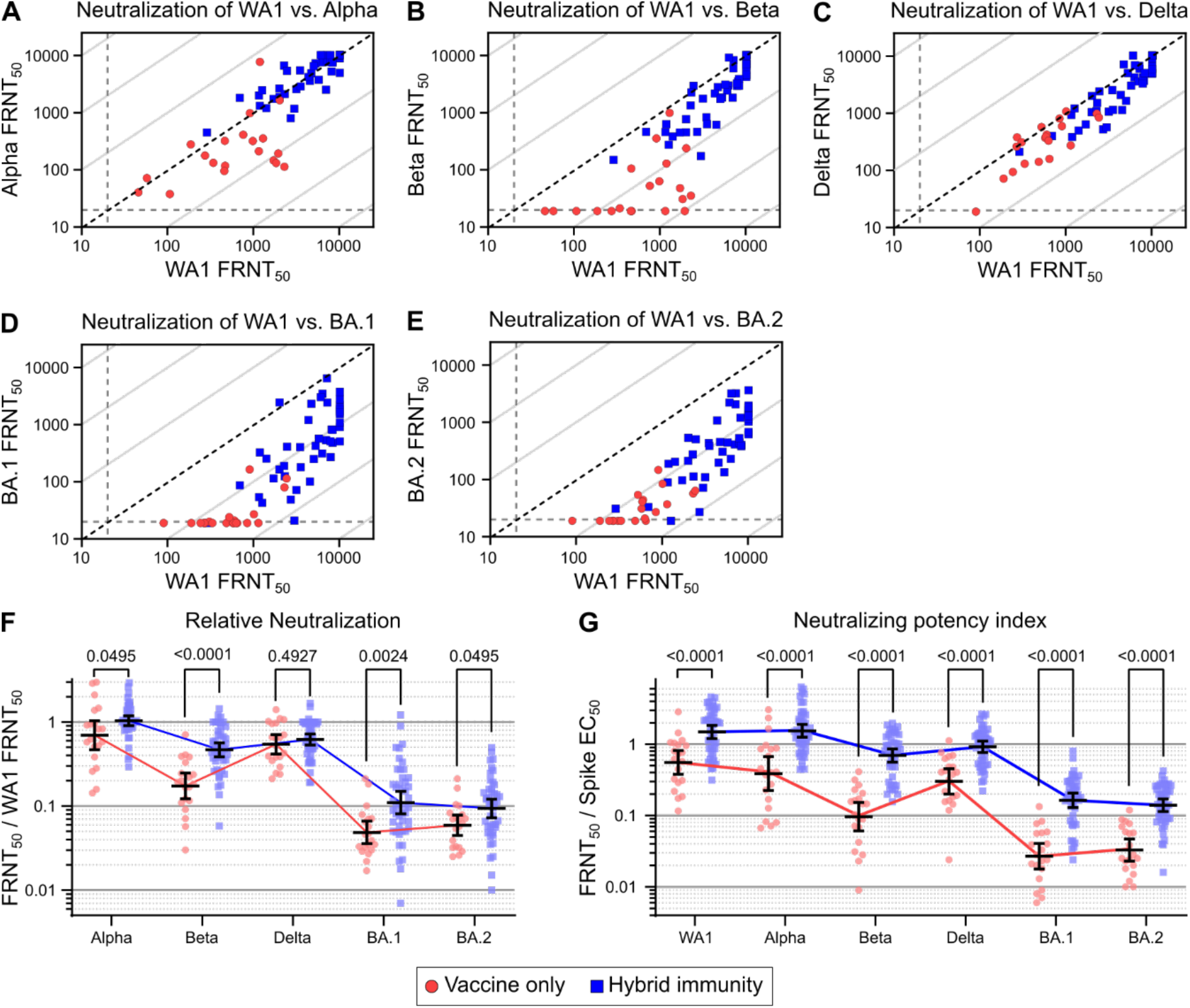
Antibody quality and variant cross-neutralization are improved with hybrid immunity. Individual neutralizing FRNT_50_ values against WA1 versus Alpha (**A**), Beta (**B**), Delta (**C**), Omicron (BA.1) (**D**), and Omicron (BA.2) (**E**). Diagonal broken line indicates equal neutralization of WA1 and variant in A-D. Relative neutralization, calculated as the neutralizing titer against each of the variants divided by the neutralizing titer against WA1 (**F**). Neutralizing potency index indicates the neutralizing FRNT_50_ against the indicated variant divided by full-length spike protein EC_50_ antibody levels (**G**). Vaccine-only participants are represented by red circles and hybrid immune participants by blue squares. Error bars represent the geometric mean with 95% confidence intervals. P values in F-G show the result of Mann-Whitney U tests with the Holm-Šídák multiple comparison correction. All P values are two-tailed and 0.05 was considered significant. For panels A-G, n=20 for the vaccine only group and n=46 for the hybrid immunity group.

To assess the potency of the neutralizing antibody responses, we then calculated the neutralizing potency index (NPI) for the individuals in each cohort against each variant. The NPI is the neutralizing titer divided by the quantity of full-length spike specific total antibody levels as measured by ELISA. NPI scores indicate the efficiency with which antigen-specific antibodies neutralize virus on a per total antibody basis in which higher scores indicate that fewer antibodies are necessary to achieve a given neutralization titer. We found that the NPI of hybrid immune individuals increased significantly for all variants tested, with indexes of 2.7-fold (WA1), 4.0-fold (Alpha), 7.2-fold (Beta), 3.0-fold (Delta), 6.1-fold (Omicron BA.1), and 4.2-fold (Omicron BA.2), indicating a significant improvement in the neutralizing efficiency of the antibodies produced by hybrid immunity compared to vaccination alone (Figure 3G).

### The interval between vaccination and natural infection dictates neutralizing titer levels

The hybrid immune cohort includes individuals who developed COVID-19 between 40 and 404 days post-vaccination, as well as individuals who were vaccinated between 35 and 283 days after testing positive for COVID-19. This range of hybrid exposure intervals allowed us to determine the impact of time intervals on the resulting neutralizing antibody response. We also characterized the correlation between antibody levels and neutralizing titers with our demographic data on age, exposure interval, sex, and the time form last exposure to sample collection. Only neutralizing antibody titers and antibody levels were significantly correlated with exposure interval. The strongest correlations were seen for full-length spike-specific antibody level, as well as neutralization of WA1, Alpha, Beta, Delta, Omicron BA.1 and Omicron BA.2 (Figure 4A-G).

**Figure 4:**
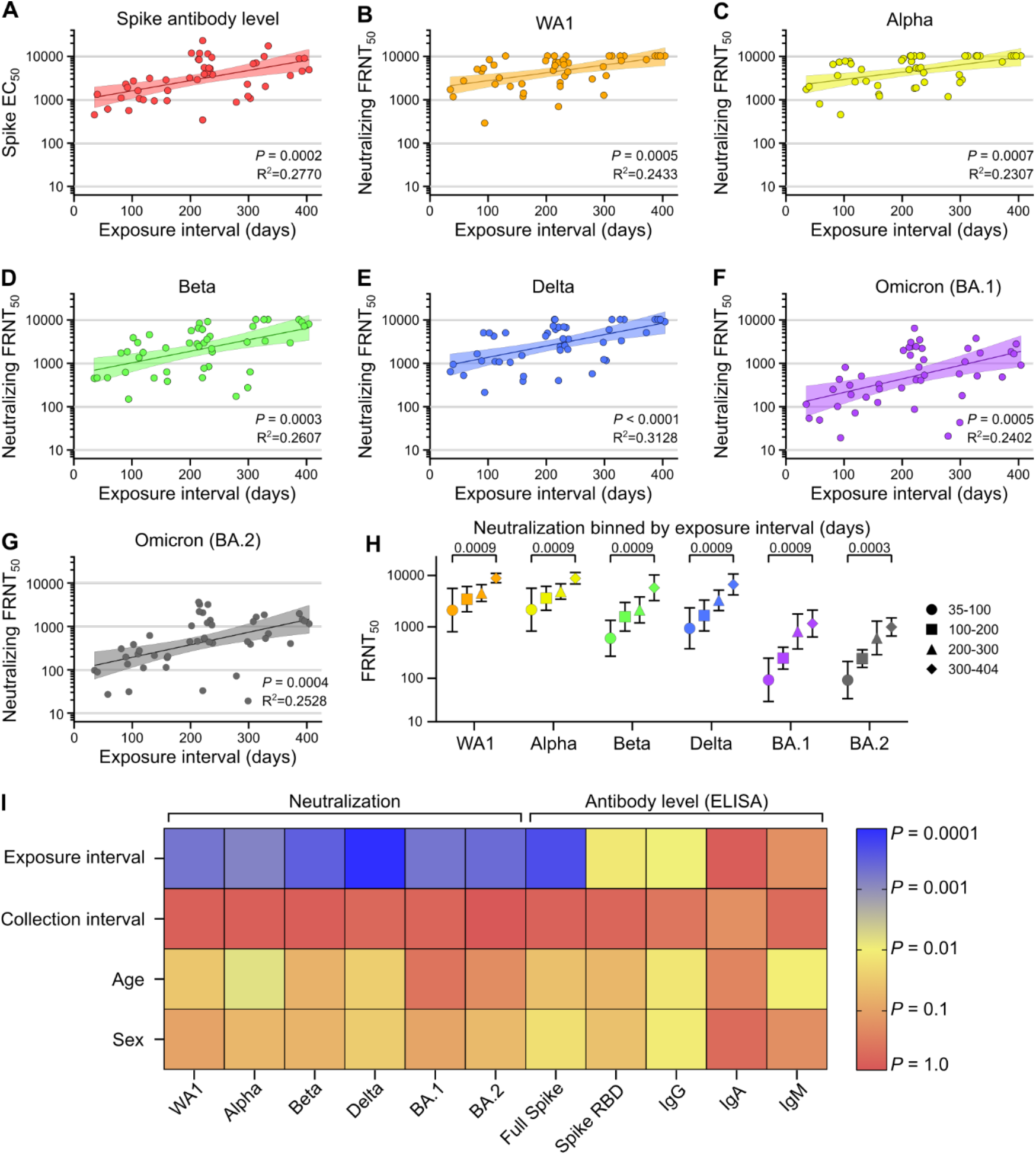
Exposure interval determines strength of hybrid immunity. Comparison of exposure interval, the time between first and last antigen exposure, with full-length spike EC_50_ antibody levels (**A**), and neutralization of WA1 (**B**), Alpha (**C**), Beta (**D**), Delta (**E**), Omicron (BA.1) (**F**), and Omicron (BA.2) (**G**). Neutralization of variants binned by exposure interval in days (**H**). Heat map of correlation significance between explanatory and response variables (**I**). Individual values in A-G are shown as filled circles and the shaded area indicates the linear fit with 95% confidence interval. R^2^ is indicated for each curve fit. P values in A-G show the result of an F-test using a zero slope null hypothesis, P values in H show the result of Mann-Whitney U tests with the Holm-Šídák multiple comparison correction, and colors in I represent the P values of pearson r correlation coefficients according to the scale bar. All P values are two-tailed and 0.05 was considered significant. For panels A-G and I, n=46. For panel H, n=7 for the 35-100 days group, n=10 for the 101-200 days group, n=18 for the 201-300 days group, and n=11 for the 301-404 days group.

The magnitude of increase seen over time was also different for each of the variants. Using linear regression, we found the neutralizing titer against WA1 increased 5.3-fold by day 400 (Figure 4). This increase was 4.8-fold for Alpha, 11.5-fold for Beta, 11.2-fold for Delta, 17.6-fold for Omicron BA.1, and 14.3-fold for Omicron BA.2. The largest increases were seen against the more contemporary variants, which also tend to be more vaccine resistant (Figure 2F). To validate that these trends are not an artifact of linear regression, we also subdivided the cohort into 100-day exposure interval bins, which recapitulated the previous findings (Figure 4H). Steady increases are seen each 100 days, resulting in a final increase of 4.2-fold against WA1, 4.1-fold against Alpha, 9.6-fold against Beta, 7.1-fold against Delta, 12.5-fold against Omicron BA.1, and 10.7-fold against Omicron BA.2 between the 35-100 and 300-404 day exposure interval groups. Both methods of analysis found a large and significant improvement in neutralizing antibody titers occurs over an increased duration between the antigen exposures provided by vaccination and natural infection. Further, these correlations were maintained when measured separately for individuals with infection prior to vaccination and individuals with vaccine breakthrough infections (supplemental figures 2 and 3). Observed separately, neutralizing titers from individuals from the breakthrough group appeared to increase faster than those in the prior infection group, but no statistically significant difference could be measured. RBD-specific total antibody and IgG levels correlated less strongly, while RBD-specific IgA and IgM did not correlate significantly with exposure interval (supplemental figure 4).

We then assessed for interactions between exposure interval and other variables that could confound our analyses, including age, sex, or the time between final antigen exposure (either vaccination or COVID-19 infection) and serum sample collection, all of which have been previously shown to affect antibody levels.^4,23,24^ As expected, titers weakly correlated with age and sex, but did not approached the relative contribution of exposure interval (Figure 4I). Collection interval was not significantly correlated with any variable, likely due to our strict 60-day limit on collection interval for inclusion in the study.

### Variant cross-neutralization improves with greater exposure intervals

After observing the improvements in variant cross-neutralization between hybrid immunity and vaccine only, we thought to determine whether there was an equivalent dependence on the exposure interval duration. Alpha is the least vaccine resistant variant and did not improve relative to WA1 because it started at a ratio of 1 from the beginning (Figure 5A). For the more vaccine resistant variants, which started well below 1, all saw increased variant cross-neutralization with increasing exposure interval (Figure 5B-E). This indicates that the neutralizing antibody response is becoming more broadly neutralizing over time, between exposures. No significant trends were seen with NPI over time (supplemental figure 5). This indicates that while the variant cross-reactivity is increasing with longer exposure intervals, the proportion of antibodies which are capable of neutralization is maintained.

**Figure 5:**
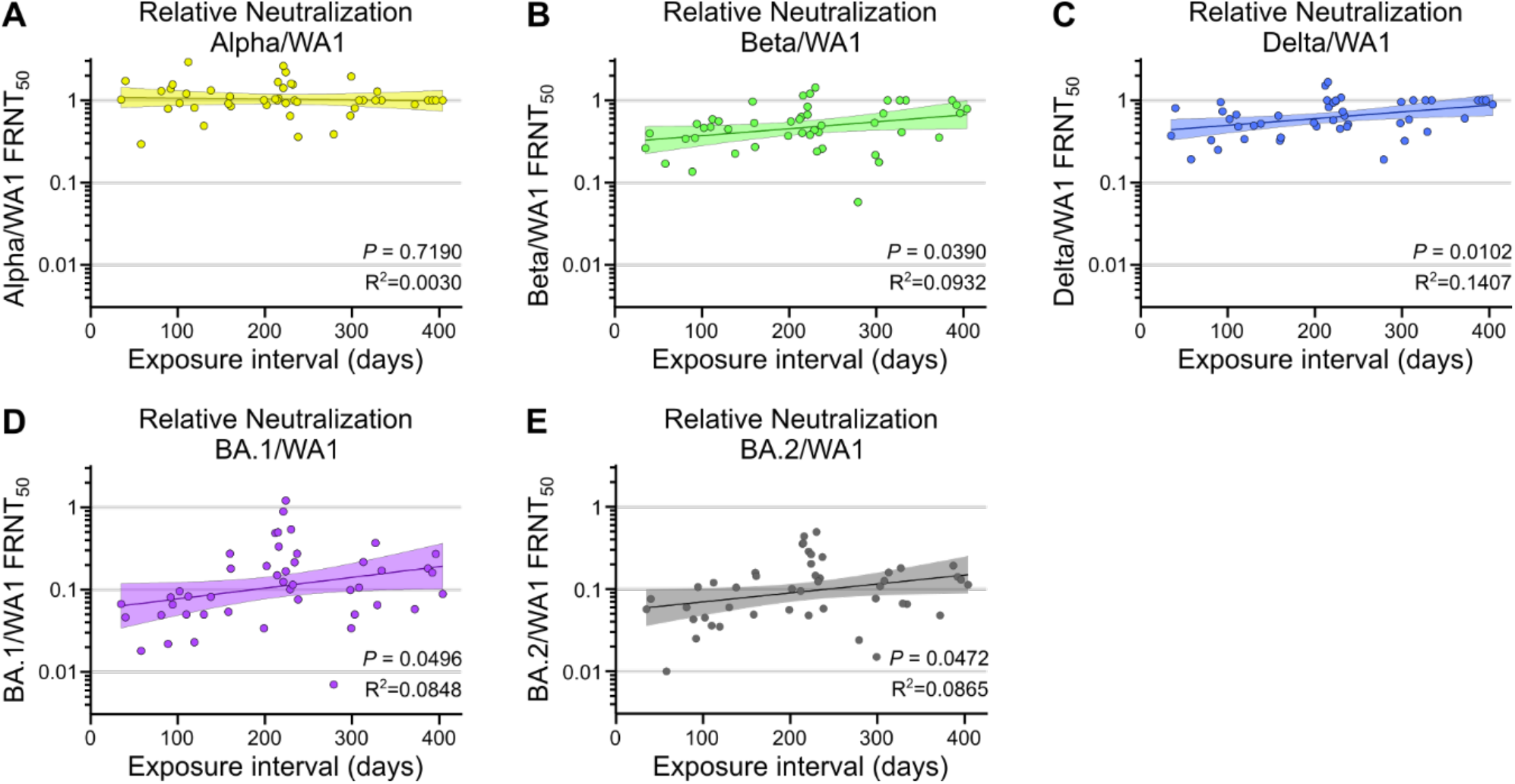
Exposure interval increases variant cross-neutralization by hybrid immune sera. Comparison of exposure interval, the time between first and last antigen exposure, with relative neutralization of Alpha (**A**), Beta (**B**), Delta (**C**), Omicron (BA.1) (**D**), and Omicron (BA.2) (**E**) over wildtype (WA1). Individual values are shown as filled circles and the shaded area indicates the linear fit with 95% confidence interval. R^2^ is indicated for each curve fit. P values show the result of an F-test using a zero slope null hypothesis. All P values are two-tailed and 0.05 was considered significant. For panels A-E, n=46.

## Discussion

This study reports superior variant-neutralizing serum antibody titers with hybrid immunity from combined vaccination and natural infection compared to vaccination alone. It further shows that longer delays, up to at least 400 days, between vaccination and infection result in the largest improvements in titers as well as better cross-neutralization of variants. The greatest increases were seen against BA.1 Omicron, which is noteworthy because the samples used in this study were collected prior to BA.1 emergence. In fact, half of the study participants were infected in the pre-vaccine era, before the emergence of any VOCs.

In our cohort, infection alone provided poor neutralizing antibody responses, while two-dose mRNA vaccination provided robust responses against original SARS-CoV-2 and the early variants, but very poor neutralization of Omicron. Hybrid immunity has been shown previously to result in greater humoral responses than two-dose vaccination,^10–13^ and our study expands upon this by identifying the hybrid exposure interval (the time between infection and vaccination) as an important factor in determining the strength of the neutralizing response. This was also recently suggested in a study of breakthrough cases over intervals up to 100 days.^19^ The finding that this effect extends to all hybrid immunity, including infection prior to vaccination is interesting because it suggests that there is nothing inherently different about the order of two different exposure modes (vaccination and infection) from the standpoint of neutralizing antibody development.

Further, because our prior infection group was never exposed to variant spike protein, it suggests that many of the conserved epitopes that the memory response develops around are present and recognizable on both the original strain of SARS-CoV-2 and every VOC including Omicron-BA.1. This hypothesis is consistent with previous work has shown that memory B cells generated by infection with original SARS-CoV-2 can recognize the variants,^25^ and that germinal center responses can continue for an extended period that improve cross-reactivity.^26–28^ Further, a recent study found that recruitment of B cells to germinal centers is controlled by the balance of existing antibody titers and availability of antigen,^29^ suggesting that antibody waning may play a direct role in broadening the antibody response over time. However, an alternative explanation is that each of the two types of hybrid immunity increase via distinct mechanisms. For instance, breakthrough infections may be more severe after longer intervals due to antibody waning in the interim, and more severe infections may lead to greater final titers. Conversely, for infection prior to vaccination, it is possible that high titers from shorter intervals result in poorer vaccine responses than at later timepoints. Neither of these alternative hypotheses explain the observation of improved variant cross-reactivity after longer intervals.

The results of this study demonstrate gradually improving memory responses to SARS-CoV-2 infection and vaccination, consistent with previous studies on the importance of an increased interval between the first two vaccine doses in achieving higher antibody levels.^17,18,20,30^ While booster vaccination has been shown to improve vaccine efficacy, there are relatively few studies that have focused on the effects of different boosting intervals.^31,32^ Currently, fourth doses are being offered to some groups in many parts of the world, and while early results are promising, it remains to be seen if continued boosting results in long-term benefits or simply a transitory bump in protective antibody levels.^33,34^

Some studies have pointed to evidence of improved durability of hybrid immune responses,^12,13,35,36^ which may be greater than that provided by boosters,^37^ but further studies are needed to establish whether vaccines which can elicit the same level of response and durability provided by hybrid immunity; perhaps the best strategy for long-term protection will involve addition of alternative vaccine types that better mimic natural infection. While hybrid immunity currently appears to offer the strongest and possibly most durable protection, intentional infection with natural COVID-19 as a means to achieve immunity is not a reasonable public health approach given the risks of severe illness, long-term complications, and death that can result from real SARS-CoV-2 infection.^38^ To the contrary, our results support increased access to vaccines. Demonstration that longer infection-vaccination intervals improve antibody responses implies that even greatly delayed vaccination will yield sizeable benefits, particularly against emerging vaccine-resistant variants. Simultaneously, our results point to a future where inevitable vaccine breakthrough infections would be expected to help build a reservoir of population-level immunity that can help blunt future waves and reduce the opportunity for further viral evolution.

## Methods

### Cohort

The longitudinal cohort participants were enrolled at Oregon Health & Science University (OHSU) immediately after receiving their first dose of the BNT162b2 COVID-19 vaccine. A pre-vaccination blood sample was collected at this time. Participants received a second vaccine dose between 20 and 32 days following the first dose, then returned between 10 and 30 days later for follow up, at which time a post-vaccination blood sample was collected.

The cross-sectional cohort was comprised of health care workers who were enrolled at OHSU, and individuals were selected from a previously established cohort based on the following criteria:^11^ Individuals who experienced COVID-19 prior to vaccination were included if serum samples were collected less than 60 days after their second vaccine dose. Vaccinated individuals who experienced vaccine breakthrough COVID-19 infections were included if serum samples were collected less than 60 days after the date of receiving a positive PCR-based COVID-19 test. Vaccinated individuals with no history of COVID-19 (vaccine only) were selected based on age, sex, days between vaccine doses, and days between final vaccine dose and sample collection in order to match the hybrid immune (combined prior infection and breakthrough) group as closely as possible.

For all participants, 4-6 mL whole blood samples were collected and then centrifuged at 1000xg for 10 minutes to isolate sera. Sera were aliquoted, heat inactivated at 65°C for 30 minutes, and frozen at -20°C until needed for laboratory tests.

### Enzyme linked Immunosorbent Assays (ELISA)

ELISA experiments were performed as previously described.^11^ Briefly, 96-well plates were coated overnight at 4°C with 1 μg/mL recombinant SARS-CoV-2 spike receptor binding domain (RBD) protein, or recombinant full-length SARS-CoV-2 spike protein. Plates were washed in phosphate buffered saline (PBS) with 0.05% Tween-20 (PBST) and blocked with PBST with 5% milk powder (dilution buffer) for one hour at room temperature (RT). Four-fold serum dilutions were prepared in dilution buffer starting at 1:50 for IgG/A/M, IgG, and IgA and 1:25 for IgM, then incubated at RT for an hour. Plates were then washed three times and incubated with secondary antibody in dilution buffer for another hour at RT. The secondary antibodies used were 1:10,000 α-IgG/A/M-HRP (Invitrogen, A18847), 1:3,000 α-IgA-HRP (Biolegend, 411002), 1:3,000 α-IgG-HRP (BD Biosciences, 555788), and 1:3,000 α-IgM-HRP (Bethyl Laboratories, A80-100P). Plates were washed three more times with PBST and developed with o-phenylenediamine (OPD) for 20 minutes then stopped with 1N HCl. Absorbance was measured at 492nm on a CLARIOstar plate reader and normalized by subtracting the average of negative control wells and dividing by the highest concentration from a positive control serum. The serum dilution that resulted in half-maximal binding was calculated by fitting normalized absorbance values to a dose-response curve as previously described,^39^ and inverse serum dilution values were reported as 50% effective concentrations (EC_50_).

### Viruses

SARS-CoV-2 clinical isolates were obtained from BEI Resources: Isolate USA-WA1/2020 [wildtype] (BEI Resources NR-52281); Isolate USA/CA_CDC_5574/2020 [Alpha - B.1.1.7] (BEI Resources NR-54011); Isolate hCoV-54 19/South Africa/KRISP-K005325/2020 [B.1.351] (BEI Resources NR-54009); Isolate hCoV-19/Japan/TY7-503/2021 [P.1] (BEI Resources NR-54982); and Isolate hCoV-19/USA/PHC658/2021 [B.1.617.2] (BEI Resources NR-55611). Isolates were propagated and titrated in Vero E6 cells as previously described.^11^ Vero E6 cells were seeded in tissue culture flasks such that they were 70-90% confluent at the time of infection. In minimal volume of Opti-MEM plus 2% FBS, flasks were infected at an MOI of 0.05 for 1 hour at 37°C before adding additional DMEM plus 10% FBS, 1% penicillin-streptomycin, 1% nonessential amino acids (complete media) to manufacturer’s recommended culture volume. Flasks were incubated until cytopathic effects were observed, 24-96 hours. Collected supernatants were centrifuged at 1,000×g for 10 minutes, aliquoted and frozen at -80°C. Titrations were performed by preparing 10-fold dilutions of frozen aliquots and incubating 30 μL for 1 hour on 96-well plates of sub-confluent Vero E6 cells before adding Opti-MEM plus 2% FBS, 1% methylcellulose (overlay media). Titration plates were incubated for 24 hours, or 48 hours for Omicron sublineages, then fixed with 4% formaldehyde for 1 hour. The formaldehyde was removed, and plates were blocked for 30 minutes at RT with PBS plus 0.1% saponin, 0.1% bovine serum albumin (perm buffer). The blocking buffer was then replaced with 1:5,000 anti-SARS-CoV-2 alpaca serum (Capralogics Inc.) in perm buffer and incubated overnight at 4°C. The plates were then washed three times for 5 minutes in PBST and incubated with 1:20,000 anti-alpaca-HRP (Novus, NB7242) for 2 hours at RT. Plates were then washed three more times with PBST for 5 minutes each, then developed with TrueBlue (SeraCare 5510-0030) for 30 minutes or until foci were strongly stained. Wells were imaged with a CTL ImmunoSpot Analyzer. Focus counts were used to calculate the concentration of focus forming units (FFU) in the virus stock aliquots.

### Focus Reduction Neutralization Test (FRNT)

Focus forming assays were performed as previously described.^11^ Briefly, Vero E6 (ATCC CRL-1586) cells were plated at 20,000 cells/well 16-24 hours before starting the assay. Sera were diluted in Opti-MEM plus 2% FBS (dilution media). Virus stocks were diluted to 3,333 FFU/mL (determined by titration) and combined 1:1 with serum dilutions. Initial serum dilutions started at 1:10, which became 1:20 after the 1:1 dilution with virus, and 30 μL of serum/virus mixture was added to each well for 1 hour at 37°C. Dilution series were performed in duplicate with one no serum control well for each replicate. Overlay media was added to each well and plates were incubated for 24 hours, or 48 hours for Omicron sublinages. Plates were fixed with 4% formaldehyde for 1 hour and then stained similarly to titration plates as described above. Foci in well images were counted with Viridot (1.0) in R (3.6.3).^40^ Percent neutralization for each well was calculated relative to the average of all no serum control wells on each plate. The serum dilution that resulted in 50% neutralization was calculated by fitting percent neutralization values to a dose-response curve as previously described,^39^ and inverse serum dilution values were reported as 50% focus reduction neutralization test (FRNT_50_) titers. For each sample, FRNT_50_ values were first calculated separately for each duplicate and verified to be within 4-fold. Combined FRNT_50_ values were calculated for all samples which passed this test, and samples which failed this test were excluded from further analysis.

### Statistical Analysis

The limit of detection (LOD) of each assay was defined by the lowest dilution tested, values below the LOD were set to LOD – 1 for both ELISA and FRNT experiments. Graphing and statistical tests were performed in GraphPad Prism. Pairwise comparisons were performed using the Mann-Whitney U test. The Holm-Šídák multiple comparison correction was used anywhere data are shown on a continuous X-axis. Simple linear regression was performed on log transformed EC_50_ and FRNT_50_ values and significance was determined with an F test with a zero-slope null hypothesis. Correlations were calculated using Pearson’s method. All P values are two-tailed and *P*=0.05 was the cutoff for significance.

### Study Approval

This study was conducted in accordance with the Oregon Health & Science University Institutional Review Board (IRB # 00022511), and written informed consent was obtained from all participants.

## Data Availability

All data produced in the present study are available upon reasonable request to the authors

## Author Contributions

TAB, HCL, WBM, MEC, FGT conceptualization. TAB, HCL, SKM, ZLL, DXL methodology. TAB software. TAB, HCL, SKM, DS, WBM, FGT, MEC validation. TAB, DS, WBM formal analysis. TAB, HCL, SKM, DS investigation. WBM, MEC, FGT resources. TAB, HCL, DS data curation. TAB writing – original draft. TAB, HCL, SKM, DS, ZLL, DXL, WBM, MEC, FGT writing – review & editing. TAB, DS visualization. TAB, WBM, MEC, FGT supervision. TAB, WBM, MEC, FGT project administration. TAB, WBM, MEC, FGT funding acquisition.

## Acknowledgements

This study was funded by a grant from the M. J. Murdock Charitable Trust (to MEC), an unrestricted grant from the OHSU Foundation (to MEC), the NIH training grant T32HL083808 (to TAB), NIH grant R011R01AI141549-01A1 (to FGT), OHSU Innovative IDEA grant 1018784 (to FGT), and NIH grant R01AI145835 (to WBM).

We thank the participants of this study for their contribution. We additionally thank the OHSU COVID-19 serology study team and the OHSU department of Occupational Health for recruiting participants and collecting samples, and the OHSU clinical laboratory under the direction of D. Hansel and X. Qin for SARS-CoV-2 testing and reporting.

## Supplementary information

**Supplemental figure 1:**
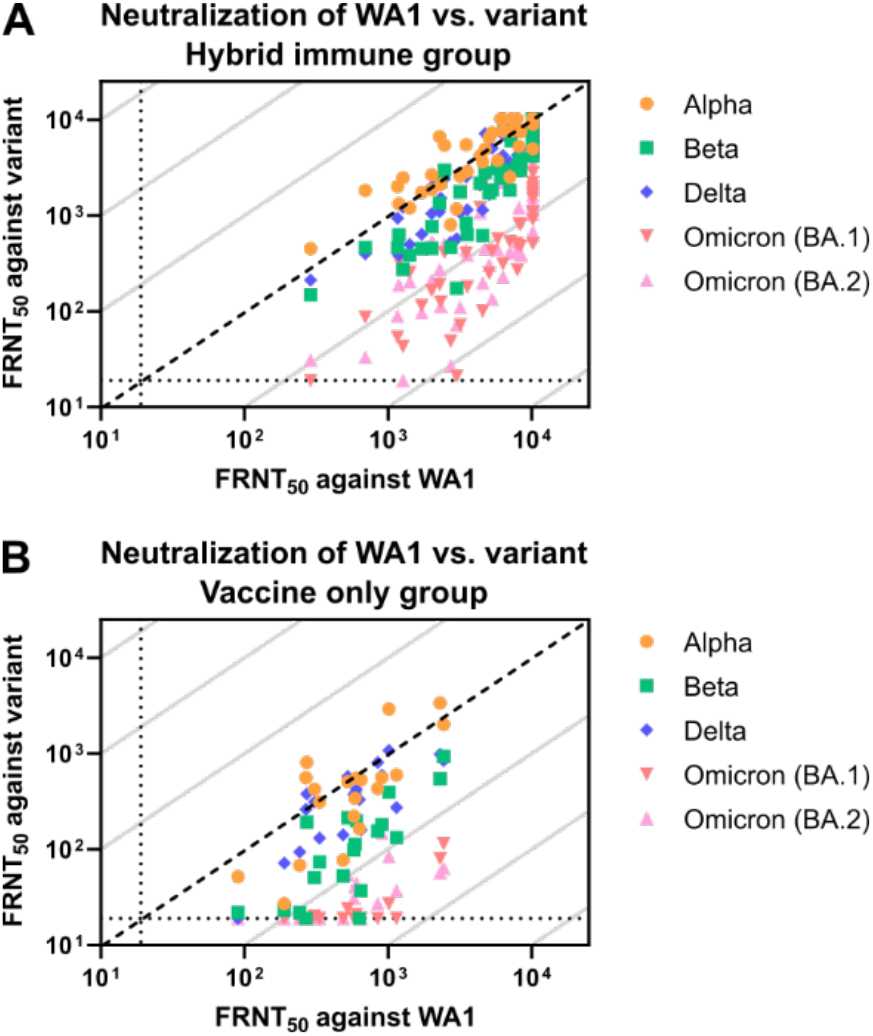
Variant cross-neutralization by hybrid immune sera is improved compared to vaccination alone. Individual neutralizing FRNT_50_ values for each of the variants against WA1 for the hybrid immune group (**A**), and two-dose vaccine only group (**B**). Diagonal broken line indicates equal neutralization of WA1 and variant. For panels A-B, n=20 for the vaccine only group and n=46 for the hybrid immunity group.

**Supplemental figure 2:**
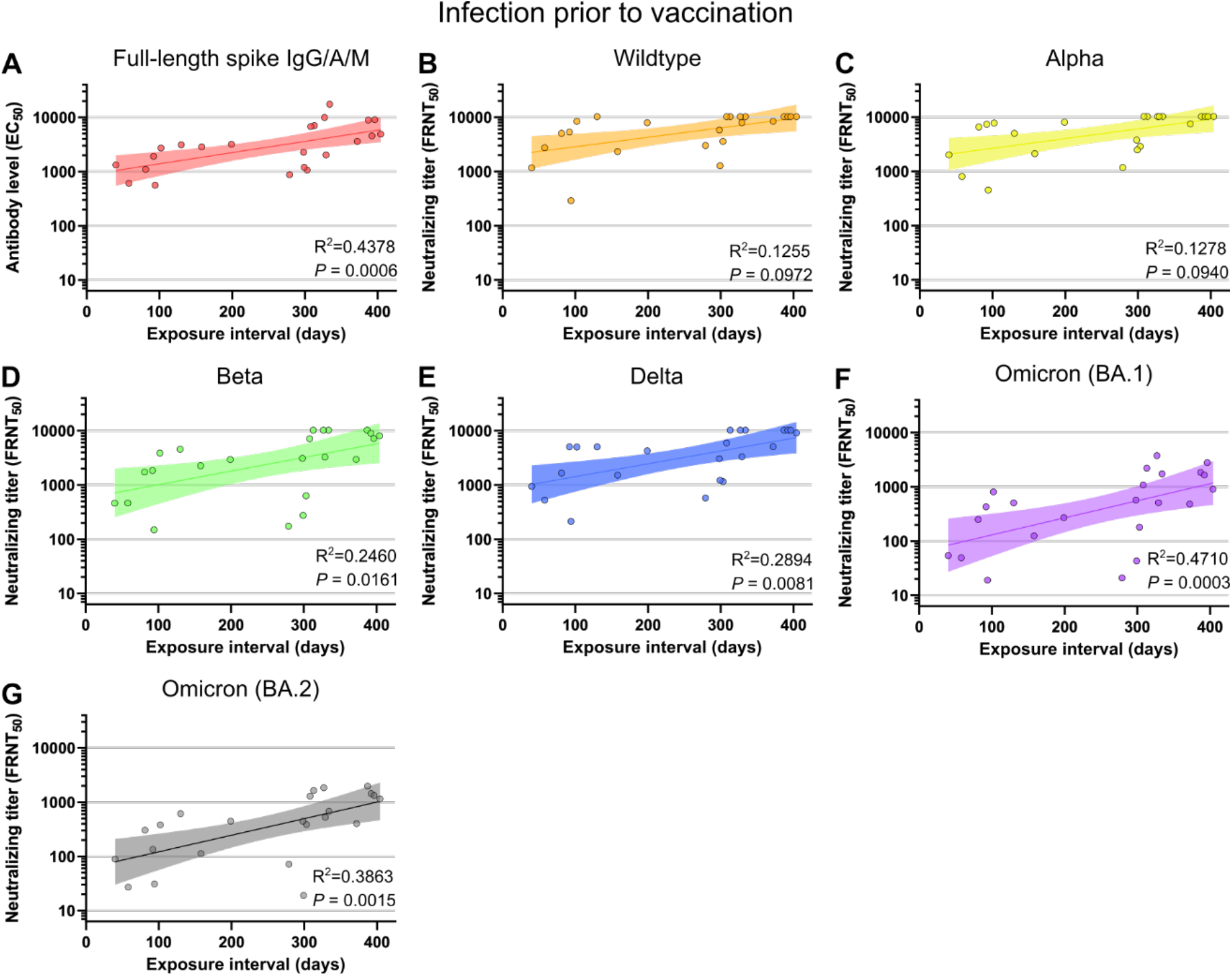
Infection prior to vaccination group neutralizing responses correlate with exposure interval. Comparison of exposure interval, the time between first and last antigen exposure, among individuals with SARS-CoV-2 infection prior to vaccination. Correlations are shown for full-length spike EC_50_ antibody levels (**A**), and neutralization of WA1 (**B**), Alpha (**C**), Beta (**D**), Delta (**E**), Omicron (BA.1) (**F**), and Omicron (BA.2) (**G**). Individual values are shown as filled circles and the shaded area indicates the linear fit with 95% confidence interval. R^2^ is indicated for each curve fit and P values show the result of an F-test using a zero slope null hypothesis. All P values are two-tailed and 0.05 was considered significant. For panels A-G, n=23.

**Supplemental figure 3:**
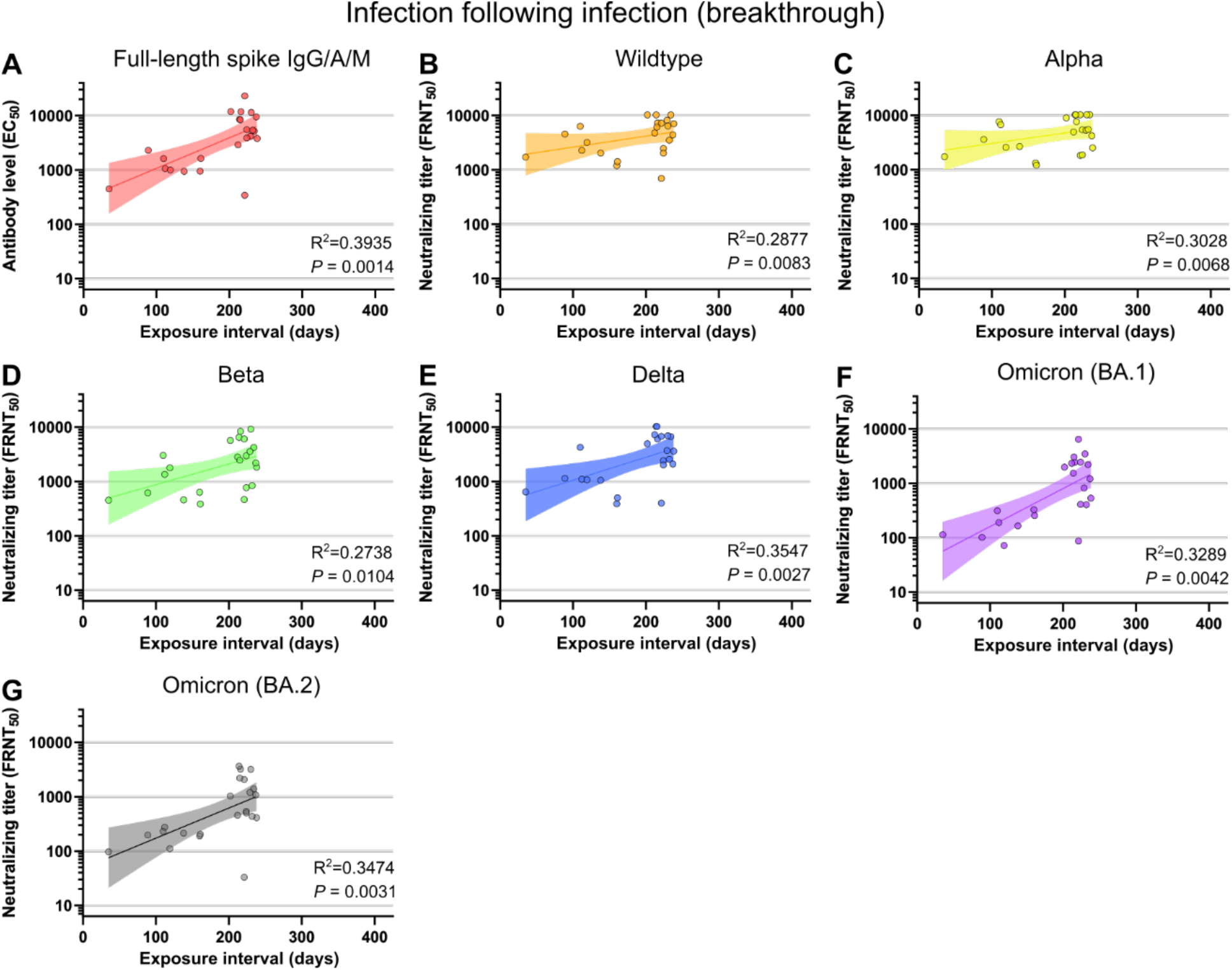
Vaccine breakthrough group neutralizing responses correlate with exposure interval. Comparison of exposure interval, the time between first and last antigen exposure, among individuals with vaccine breakthrough infections. Correlations are shown for full-length spike EC_50_ antibody levels (**A**), and neutralization of WA1 (**B**), Alpha (**C**), Beta (**D**), Delta (**E**), Omicron (BA.1) (**F**), and Omicron (BA.2) (**G**). Individual values are shown as filled circles and the shaded area indicates the linear fit with 95% confidence interval. R^2^ is indicated for each curve fit and P values show the result of an F-test using a zero slope null hypothesis. All P values are two-tailed and 0.05 was considered significant. For panels A-G, n=23.

**Supplemental figure 4:**
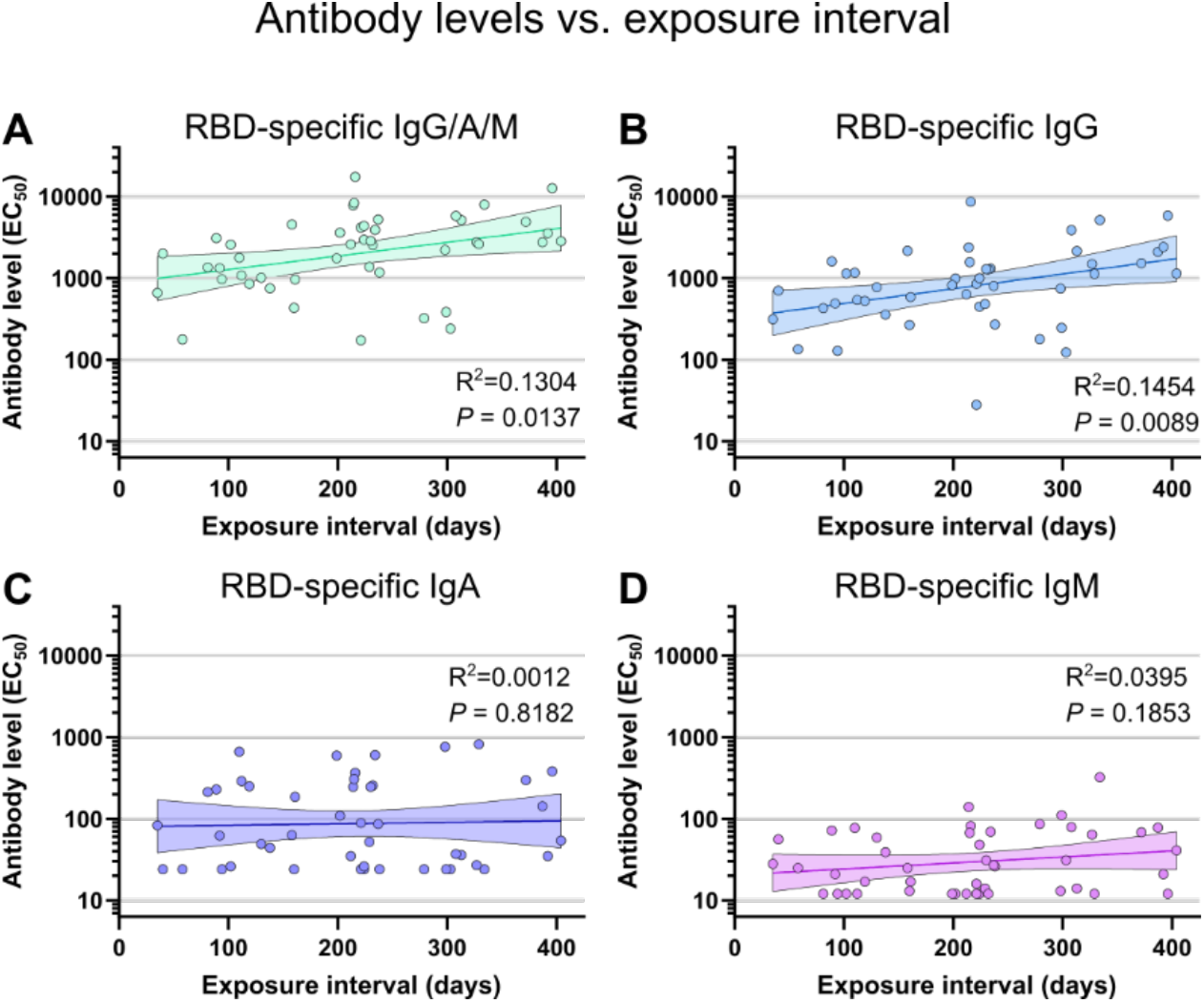
Other antibody isotypes correlate less well with exposure interval. Comparison of exposure interval, the time between first and last antigen exposure, with total (IgG/A/M) spike RBD (**A**), IgG (**B**), IgA (**C**), and IgM (**D**) EC_50_ antibody levels. Individual values are shown as filled circles and the shaded areas indicate the linear fit with 95% confidence interval. R^2^ is indicated for each curve fit and P values show the result of an F-test using a zero slope null hypothesis. All P values are two-tailed and 0.05 was considered significant. For panels A-D, n=46.

**Supplemental figure 5:**
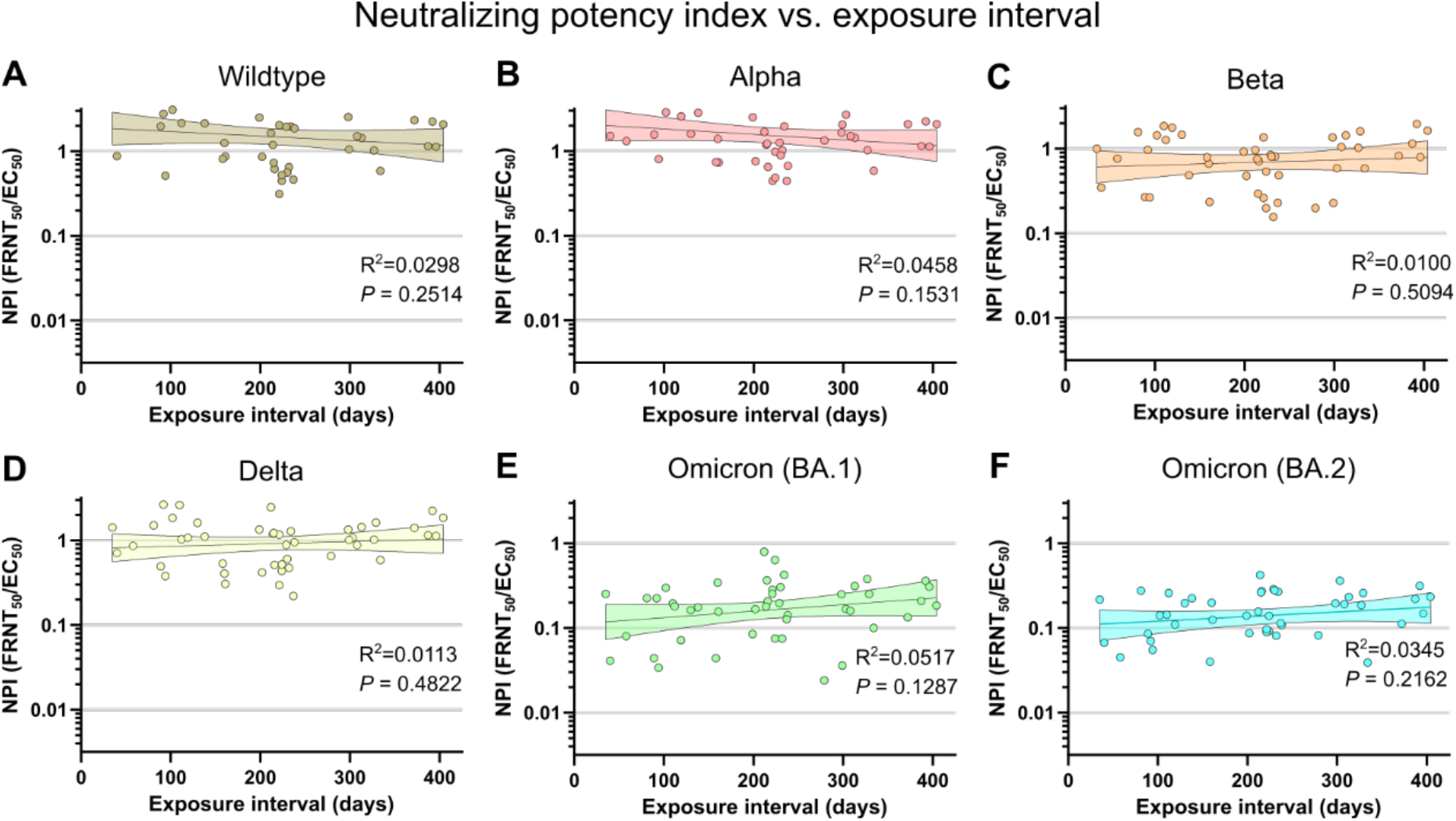
Neutralizing potency index does not correlate with exposure interval. Comparison of exposure interval, the time between first and last antigen exposure, with neutralization potency index (FRNT_50_ / full-length spike EC_50_) of wildtype (WA1) (**A**), Alpha (**B**), Beta (**C**), Delta (**D**), Omicron (BA.1) (**E**), and Omicron (BA.2) (**F**). Individual values are shown as filled circles and the shaded area indicates the linear fit with 95% confidence interval. R^2^ is indicated for each curve fit. P values show the result of an F-test using a zero slope null hypothesis. All P values are two-tailed and 0.05 was considered significant. For panels A-F, n=46.

